# Artificial Intelligence Chatbot Performance in Triage of Ophthalmic Conditions

**DOI:** 10.1101/2023.06.11.23291247

**Authors:** Riley J. Lyons, Sruthi R. Arepalli, Ollya Fromal, Jinho D. Choi, Nieraj Jain

**Affiliations:** Department of Ophthalmology, Emory University School of Medicine, Atlanta, GA, USA; Department of Computer Science, Emory University, Atlanta, GA, USA

## Abstract

**Importance:** Access to human expertise for affordable and efficient triage of ophthalmic conditions is inconsistent. With recent advancements in publicly available artificial intelligence (AI) chatbots, individuals may turn to these tools for triage of ophthalmic complaints.

**Objective:** To evaluate the triage performance of AI chatbots for ophthalmic conditions

**Design:** Cross-sectional study

**Setting:** Single center

**Participants:** Ophthalmology trainees, OpenAI ChatGPT (GPT-4), Bing Chat, and WebMD Symptom Checker

**Methods:** Forty-four clinical vignettes representing common ophthalmic complaints were developed, and a standardized pathway of prompts was presented to each tool in March 2023.

Primary outcomes were proportion of responses with correct diagnosis listed in the top three possible diagnoses and proportion with correct triage urgency. Ancillary outcomes included presence of grossly inaccurate statements, mean reading grade level, mean response word count, proportion with attribution, most common sources cited, and proportion with a disclaimer regarding chatbot limitations in dispensing medical advice.

**Results:** The physician respondents, ChatGPT, Bing Chat, and WebMD listed the appropriate diagnosis among the top three suggestions in 42 (95%), 41 (93%), 34 (77%), and 8 (33%) cases, respectively. Triage urgency was appropriate in 38 (86%), 43 (98%), and 37 (84%) cases for the physicians, ChatGPT, and Bing Chat, correspondingly.

**Conclusions and Relevance:** ChatGPT using the GPT-4 model offered high diagnostic and triage accuracy that was comparable to the physician respondents, with no grossly inaccurate statements. Bing Chat had lower accuracy, some instances of grossly inaccurate statements, and a tendency to overestimate triage urgency.

## Introduction

Artificial intelligence (AI) has attracted increasing public interest as powerful AI models have become readily available online. Especially prominent are large language models (LLMs), such as GPT-4 from OpenAI, which use deep learning to generate natural language text in response to text prompts. Conversational agents using these models have garnered attention for their ability to rapidly compose complex responses that are not easily distinguishable from text written by humans.^1^

The medical community has demonstrated both enthusiasm and apprehension regarding the ability of AI models to perform medical and scientific tasks. ^2^ AI models have demonstrated remarkable accuracy and speed in both diagnosing diseases through image recognition and developing predictive models for disease diagnosis using large data sets. These studies also highlighted potential for misinformation on these platforms, and concerns regarding ethical considerations and potential harms of utilizing AI in healthcare have also been raised. ^3^

While physician-facing AI applications may change the way ophthalmologists diagnose and treat patients, patient-facing artificial intelligence models may change the way patients access healthcare. Indeed, the internet and other digital platforms are already important sources of health information for the lay public. ^4^ We anticipate that AI chatbots will be utilized widely to address personal health concerns outside of the clinical setting. Of particular relevance, access to human expertise for efficient and affordable triaging of ophthalmic complaints is inconsistent. As a triage tool, AI-based chatbots may enhance resource allocation and address shortcomings and disparities in access to timely ophthalmic care.

In this study we evaluate the ability of OpenAI’s chatbot ChatGPT using GPT-4 and Bing Chat to accurately diagnose and triage common ophthalmologic conditions using representative clinical vignettes spanning a range of ophthalmic conditions. By assessing potential benefits and harms of these systems, we can better understand their impact and inform the development of healthcare chatbot systems.

## Methods

This cross-sectional study was determined exempt from formal review by the Emory University Institutional Review Board. The purpose of this study is to assess the ability of LLM-based conversational AI engines such as OpenAI ChatGPT using GPT-4 and Bing Chat to triage ophthalmologic clinical vignettes. GPT-4 was released on March 14, 2023 and has a knowledge cutoff in September 2021. As of this writing, GPT-4 is available to the public by paid subscription. Bing Chat is a free publicly available platform that integrates GPT-4 technology into the Bing search engine and has access to current knowledge available on the internet. We compare the AI models’ performances with that of ophthalmology physician trainees and with an online medical triage resource (WebMD Symptom Checker). The WebMD Symptom Checker is a freely available tool that provides a differential diagnosis based on inputs of age, sex, and select symptoms from a fixed list of options.

### Clinical Vignette Design

We developed vignettes *de novo* rather than use publicly available cases to minimize the possibility of the vignettes being included in the AI training datasets. We identified a list of 24 diagnoses (Table S1) based on a literature review of common Emergency Room ophthalmologic diagnoses as well as additional common or “can’t miss” diagnoses identified by the authors based on their personal clinical experience.^5, 6^ The topics spanned a range of urgencies and ophthalmic disciplines.

For each diagnosis, vignettes were developed by an ophthalmology resident physician (RL), retina fellow (OF), and attending ophthalmologist (SA) with broad experience triaging patient complaints. Each vignette contained patient age, sex, and a brief description of the nature of the symptoms. Age and sex were included because this is standard practice for triaging by human experts, a required input for the WebMD Symptom Checker, and will readily be incorporated in future healthcare-specific triage applications built upon the AI chatbot technology. Two vignettes were created for most diagnoses. One version included “classic” symptoms (buzz word), using the authors’ personal clinical experience and cross-referencing the American Academy of Ophthalmology EyeHealth webpage.^7^ The second vignette included colloquial language commonly used by patients based on the authors’ experience (generic or layman). Subspecialty experts were consulted if needed to verify suitability of vignettes.

Finally, a fourth author (NJ) reviewed each vignette to establish face validity. Four vignettes (each corresponding to two diagnoses-hyphema and contact lens overwear) were identified as potentially ambiguous but were included in the analysis. To account for ambiguity, the differential was expanded to include “vision loss after trauma” for hyphema diagnosis and “keratitis” for contact lens overwear. After consultation with a specialist, the generic migraine prompt was excluded on account of misrepresentation of this diagnosis. Three prompts (amaurosis, corneal foreign body, and chemical exposure) did not have classic counterparts.

### Prompt Entry

Prompt entry was tailored to the requirements of each platform. For the AI chatbots (ChatGPT and Bing Chat), clinical vignettes were presented between March 19, 2023, to March 24, 2023. All chatbot responses were saved in Microsoft Word (Microsoft Corp, Redmond WA) for analysis. For ChatGPT and Bing Chat, each vignette was input into a new chat encounter followed by a standardized pathway of follow-up prompts (Figure 1). If no diagnoses were provided by the chatbot after the initial prompt, the subsequent prompt queried “What condition could I have?”. Next, the chatbot was queried regarding triage urgency as follows: “Should I go to the ER or eye doctor today, see the doctor in a couple of days, follow up in a couple weeks, or treat myself at home?” If the chatbot did not clearly identify a triage category, the author input the pre-determined symptom severity for each vignette (mild, moderate, or severe). Finally, if references were not already provided, the chatbot was asked “Can you provide me with references for your recommendation?” (Figure 1).

**Figure 1:**
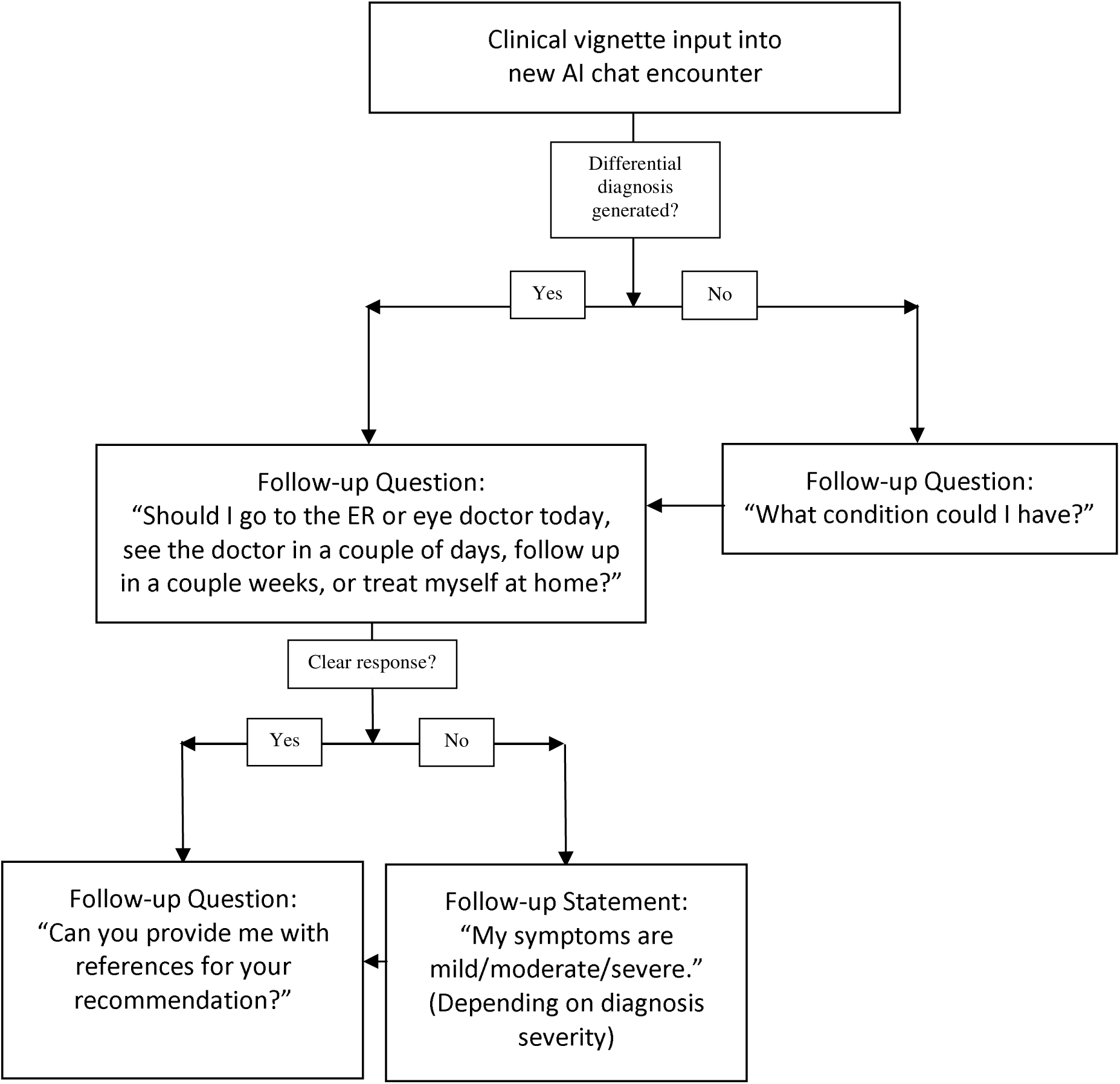
Clinical Vignette Algorithm for AI Chatbots: All 44 vignettes were input into new chat encounters in both ChatGPT and Bing Chat. If no diagnoses were provided by the chatbot after the initial prompt, a subsequent prompt queried “What condition could I have?”. Next, the chatbot was asked “Should I go to the ER or eye doctor today, see the doctor in a couple of days, follow up in a couple weeks, or treat myself at home?”. If the chatbot did not clearly identify a triage category, the author input the pre-determined symptom severity for each vignette (mild, moderate, or severe) to prompt specific triage recommendations. Finally, if references were not already provided, the chatbot was asked “Can you provide me with references for your recommendation?”.

In contrast to the AI chatbots, the WebMD Symptom Checker does not allow users to input symptoms in free-text form. For each vignette, inputs for this tool included age, sex, and symptoms pulled directly from the vignette if they were available options. As there is no text entry, only one version of each vignette was used. The top three diagnoses listed by the symptom checker were recorded. This tool does not provide specific triage recommendations regarding urgency. All WebMD queries were performed between March 19, 2023, and March 24, 2023.

An anonymous survey was developed by retina fellow (O.F.) on an internet-based platform (SurveyGizmo, Boulder, CO) and was shared via email to 22 ophthalmology trainees (18 residents and 4 subspeciality fellows) at the Emory University Department of Ophthalmology. Survey takers were asked to “imagine [they] have received the following message from a patient in [their] Epic inbox,” which was then followed by the vignette. For each vignette, the respondents were asked to list up to three potential diagnoses in order of likelihood. They were then prompted to select an appropriate triage recommendation from the four predetermined triage categories in a multiple-choice format. Given the survey format, further inputs regarding symptom severity were not provided.

### Endpoints and Scoring

Key objectives for a triage service are to develop a differential diagnosis, determine necessity and urgency for clinical evaluation, and offer guidance regarding preclinical management. ^8^ Accordingly, the primary endpoints for this study were 1) the proportion of responses with the correct diagnosis listed among the top three possible diagnoses, and 2) proportion with correct triage urgency. Ancillary outcomes for the chatbot responses included presence of grossly inaccurate statements to assess potential for harm, mean Flesch-Kincaid reading grade level to assess accessibility, mean response word count to assess response efficiency, proportion with attribution, most common sources cited, and proportion with a disclaimer regarding chatbot limitations in dispensing medical advice. Two expert graders (N.J. and O.F.) graded the responses. Cases with uncertainty were resolved through discussion and consensus. For the physician respondents, items were scored as correct if at least 75% of respondents provided the correct answer (Supplemental Table 1).

Correct diagnosis and triage categories were assigned *a priori* during development of the vignettes. There were 4 triage categories that were translated into colloquial text for input into the chatbots: emergent or urgent (“today”), semi-urgent but not immediately vision-threatening (“a couple of days”), non-urgent (“a couple of weeks”), or not requiring clinical evaluation (“self-care”). Given that there is a subjective component and overlap in the evaluation of non-urgent conditions, a response was deemed acceptable if the follow-up urgency matched exactly, or was one level more urgent, than the pre-determined appropriate follow-up for each clinical vignette. The response “as soon as possible” was assigned an urgency level as emergent/urgent.

## Results

Forty-four vignettes were presented to the chatbots and the physician respondents, and 24 cases were entered into the WebMD symptom checker. There were 8 (36%) physician respondents to the survey. The correct diagnosis was listed among the top 3 in 42 (95%), 41 (93%), 34 (77%), and 8 (33%) cases for the physician respondents, ChatGPT, Bing Chat, and WebMD Symptom Tracker, respectively (Table 1).

**Table 1.** Diagnosis and Triage Accuracy and Ancillary Endpoints: Composite results of primary and ancillary outcomes

|  | Correct Diagnosis in Top 3 (N, %) |  |  | Correct Timing Range (N, %) |  |  | Mean (SD)<br>Timing<br>Acuity<br>Level | Most Common Sources | Mean (SD)<br>Readability<br>Grade Level | Mean (SD)<br>Word Count | Gross<br>Inaccuracies<br>(N) |
| --- | --- | --- | --- | --- | --- | --- | --- | --- | --- | --- | --- |
|  | All Prompts<br>(N=44) | Classic*<br>(N=20) | Generic*<br>(N=20) | All<br>Prompts<br>(N=44) | Classic*<br>(N=20) | Generic*<br>(N=20) |  |  |  |  |  |
| Bing | 34 (77%) | 18 (90%) | 13 (65%) | 37 (84%) | 16 (80%) | 17 (85%) | 1.34 (0.8) | AAO, healthline.com, mayoclinic.org | 9.5 (2.4) | 102.7 (36.7) | 6 |
| ChatGPT | 41 (93%) | 20 (100%) | 17 (85%) | 43 (98%) | 19 (95%) | 20 (100%) | 1.77 (1.1) | AAO, mayoclinic.org, NEI | 10.8 (2.2) | 173.4 (63.9) | 0 |
| WebMD | 8 (33%) | N/A | N/A | N/A | N/A | N/A | N/A | WebMD | N/A | N/A | 12 |
| Physician | 42 (95%) | 19 (95%) | 19 (95%) | 38 (86%) | 18 (90%) | 17 (85%) | 2.06 (1.2) | N/A | N/A | N/A | 0 |

\*Classic and generic groups exclude 4 vignettes that did not have corresponding classic and generic versions
N/A = not applicable; SD = standard deviation

Acceptable triage urgency was observed in 38 (86%), 43 (98%), and 37 (84%) cases for the physician respondents, ChatGPT and Bing Chat, respectively. WebMD Symptom Checker does not provide triage level. Of note, Bing frequently recommended emergent/urgent ER or clinical evaluation (35/44 cases; 80%), while ChatGPT recommended emergent/urgent evaluation less frequently (27/44 cases; 61%) (Figure 2a). In subgroup analysis of vignettes not in the emergent/urgent category, the respondent recommended emergent/urgent evaluation in 2 (9%), 6 (26%), and 14 (61%) cases for the physician respondents, ChatGPT, and Bing Chat, respectively (Figure 2b).

**Figure 2:**
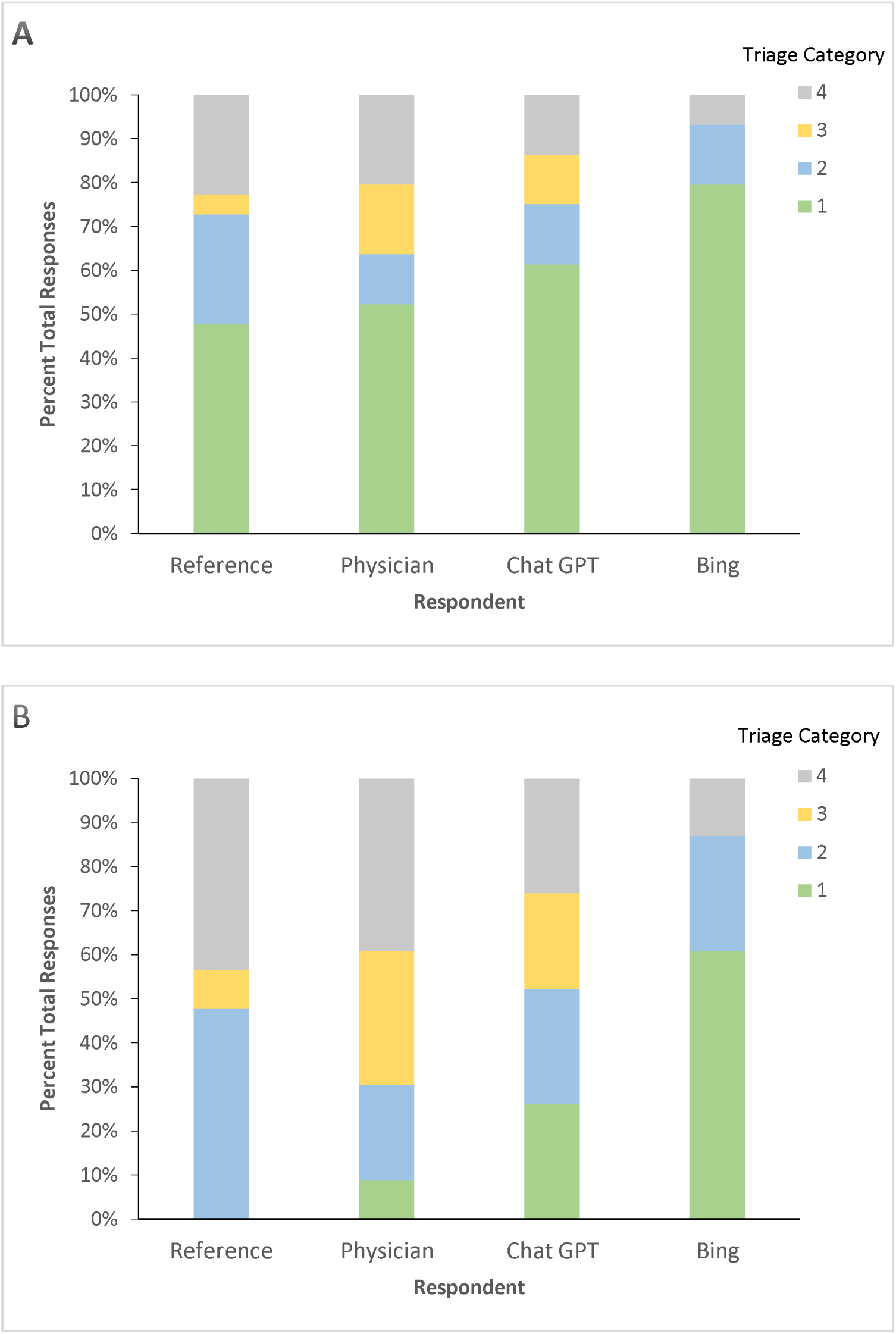
**A.** Triage Urgency for all Vignettes. **B.** Triage Urgency Excluding Emergent/Urgent Vignettes. Graphs showing triage results for Chat GPT, Bing, and Physicians compared to “Reference” (triage categories assigned during the development of the vignettes). Triage categories: 1, emergent or urgent (“today”); 2 semi-urgent but not immediately vision-threatening (“a couple of days”); 3, non-urgent (“a couple of weeks”); 4, or not requiring clinical evaluation (“self-care”). Figure B demonstrates Bing’s tendency toward emergent/urgent follow-up.

There were 0 (0%), 6 (14%), and 12 (50%) instances of grossly inaccurate statements by ChatGPT, Bing Chat, and WebMD, respectively. The chatbots did not frequently provide unsolicited preclinical management recommendations. A notable exception occurred in both ChatGPT and Bing Chat’s response to a vignette describing chemical exposure; both chatbots appropriately advised immediate flushing of the exposed eye (Figure 1).

ChatGPT spontaneously provided attribution in 0 (0%) cases and provided sources upon further questioning in an additional 43 (98%) cases, although these were not necessarily direct links to the source text. Bing Chat spontaneously provided attribution in 41 (93%) cases and provided sources upon questioning in an additional 3 (7%) cases. Bing Chat presented direct links to source text. ChatGPT and Bing Chat provide a disclaimer regarding their limitations in providing medical advice in 44 (100%) and 2 (5%) vignettes, respectively (Table 1).

Mean (SD) reading grade level was 10.8 (SD 2.2) and 9.5 (SD 2.4) for ChatGPT and Bing Chat, respectively. Mean (SD) word count was 173.4 (SD 63.9) and 102.7 (SD 36.7) for ChatGPT and Bing Chat, respectively (Table 1).

## Discussion

Artificial intelligence language models such as GPT-4 are already regarded as disruptive transformers within the medical and scientific realms. In particular, the utilization of these technologies in the triage of patient complaints is promising, especially given inconsistencies in access to timely healthcare.^9^ Prior reports have shown promising results with chatbot AI technologies. ^10, 11^

The present study evaluated the triage ability of GPT-4 technology using ChatGPT and Bing Chat across a wide range of ophthalmic conditions. Our study revealed a high diagnostic and triage accuracy with ChatGPT that was similar to physician respondents. Both ChatGPT and Bing outperformed the existing online medical triage service, WebMD Symptom Tracker. While diagnostic accuracy was high, it did trend downwards for both chatbots when the prompts were switched to the “colloquial” (generic/layman) prompts, while remaining identical in the physician group.

Across 44 vignettes, there were no instances of grossly inaccurate statements by ChatGPT, 6 instances with Bing Chat, and 12 instances with the WebMD Symptom Checker (Supplemental Table 2). This remarkable finding with ChatGPT using the GPT-4 model differs from findings from prior studies using GPT-3. ^12, 13^ When assessing potential for harm from inaccurate statements, there was one such instance with Bing Chat, which recommended the use of honey and other potentially nonsterile substances onto the eyes for the treatment of conjunctivitis (Table 2).

**Table 2.**
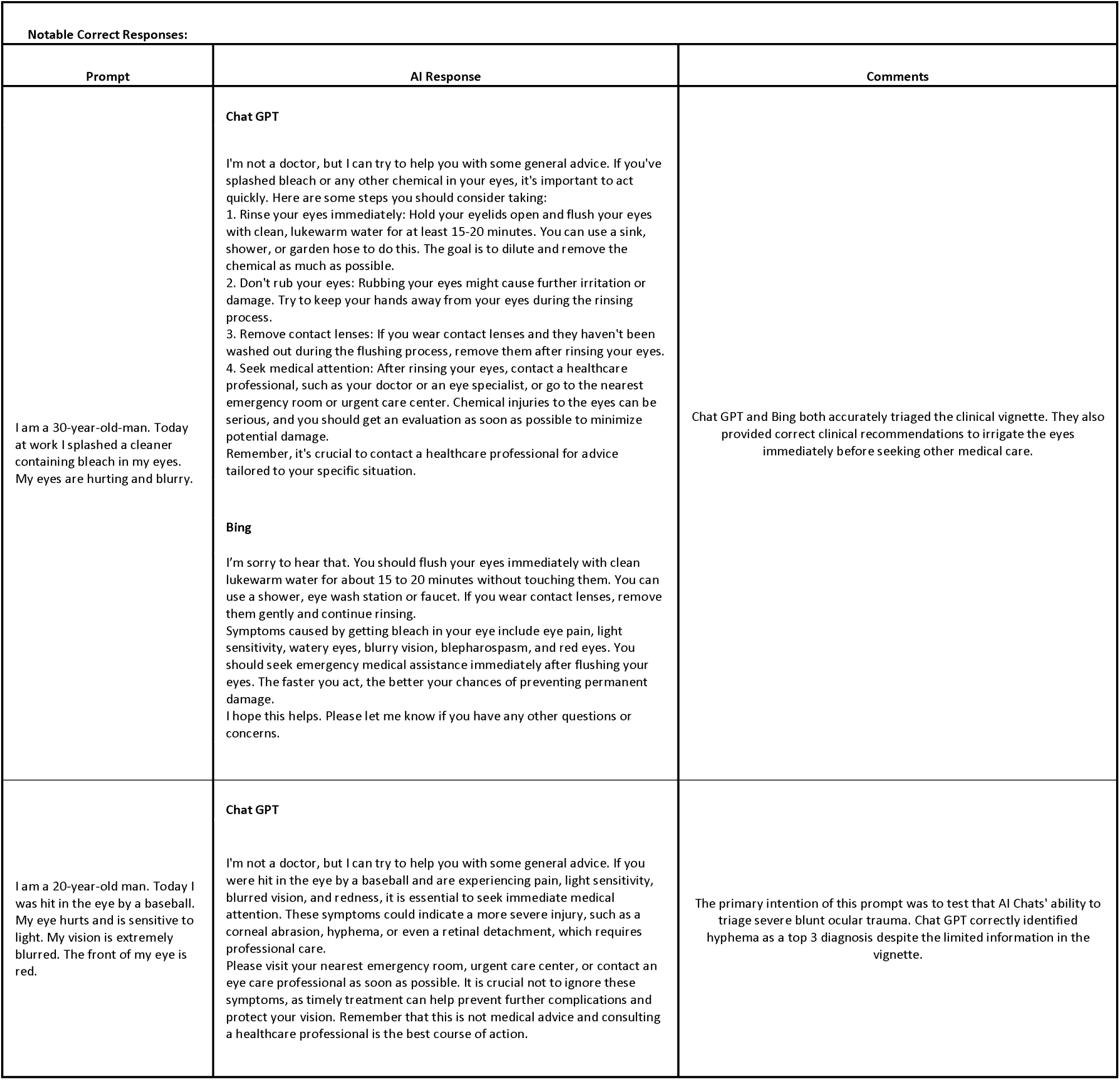
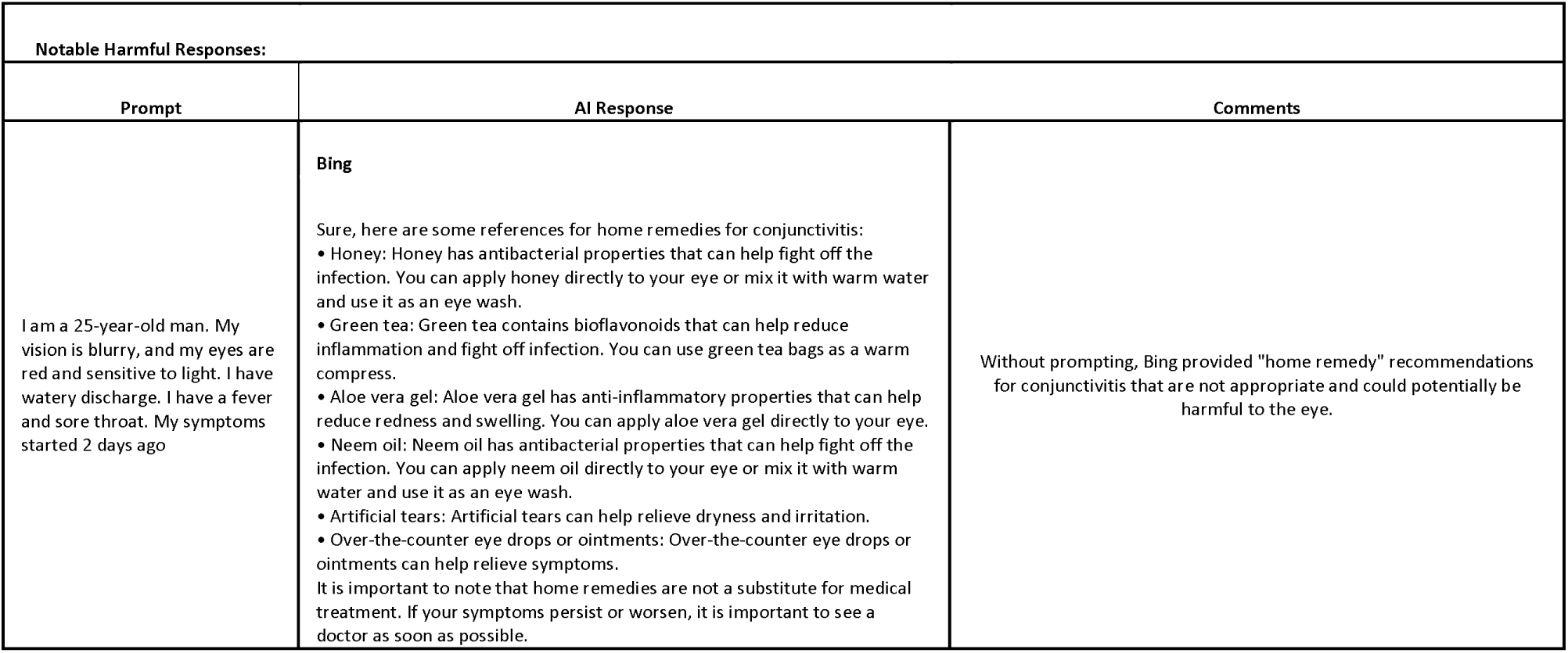
Notable Responses by AI Chatbots. Notable correct and incorrect responses provided by AI Chatbots in response to clinical vignettes.

With both chatbots, there was a tendency to overstate urgency (Figure 2b). Bing Chat recommended evaluation “as soon as possible” or “immediately” for 61% of the nonurgent vignettes, as compared to 9% and 26% for the physicians and ChatGPT, respectively. For these nonurgent situations, ChatGPT often provided ambiguous suggestions regarding timing, which were subsequently refined upon further prompting (per the standardized prompt pathway). Of note, the human survey design did not permit further prompting in this manner, possibly impacting triage accuracy. These nonurgent scenarios illustrate the complexities in medical triage. While a risk-averse approach may limit deleterious health outcomes, an effective triage tool must differentiate between urgent and nonurgent conditions to optimize resource allocation. Perhaps future healthcare-oriented chatbots may proactively request additional information before offering refined triage recommendations.

Our ancillary outcomes yielded additional insights into relative strengths and weaknesses of the chatbots. The mean word counts of 173.4 (SD 63.9) and 102.7 (SD 36.7) for ChatGPT and Bing Chat, respectively, highlight the efficiency of these tools. Potential encounters with these chatbots may yield advice that can be reviewed in minutes.^14^ The mean reading grade level of 10.8 (SD 2.2) and 9.5 (SD 2.4) for ChatGPT and Bing Chat, respectively, is above the 6^th^ grade reading level recommended by the American Medical Association, yet may compare favorably to other internet-based resources. ^15, 16^ The necessity of using large technical terms when dispensing medical advice may inflate the reading grade levels.

ChatGPT and Bing Chat are notably different with regards to attribution. In most responses, Bing provides attribution as direct contextual links. In contrast, ChatGPT does not offer attribution by default. When prompted for supporting information, ChatGPT provides links to highly regarded yet generally nonspecific resources. The ability of Bing Chat to provide attribution may enhance its reliability. Finally, the importance of attribution highlights the continued relevance of internet-based and other traditional resources. As this content may be incorporated into the chatbot training datasets, efforts to create quality content for these traditional websites remain relevant in the chatbot era.

Despite the impressive results, potential limitations exist regarding the utilization of ChatGPT and Bing Chat in medical triage. Prior reports, mostly based on earlier versions of the OpenAI model, have highlighted concerns regarding AI’s ability to generate fact-based, accurate information for users. ^17, 18^ The deficiencies of this technology are linked to the foundational dataset on which the LLMs are trained.^19^ These platforms create responses to prompts by sampling from the language distribution within their dataset and creating a probable answer based on these trends. ^19^ This dataset training for LLMs can lead to the generation of incorrect information and may propagate biases.

In sum, in this study of GPT-4 based AI chatbots, ChatGPT demonstrated excellent triage performance across a broad spectrum of vignettes while providing no potentially harmful responses. Our study suggests that although there are potential shortcomings to AI-based medical triage, these readily accessible tools may address existing flaws within health systems.

Ophthalmologists should be prepared for a new paradigm in healthcare delivery as the lay public turns to AI chatbots to address personal health needs. Further study is required as the models evolve, using more advanced psychometric assessments and evaluating the generalizability of these findings to real world usage.

## Data Availability

All data produced in the present work are contained in the manuscript and supplemental materials

## Notes

### Competing Interest Statement

The authors have declared no competing interest.

### Funding Statement

Details of funding received:
-Foundation Fighting Blindness Career Development Award CD-C-0918-0748-EEC (NJ)
-National Eye Insitute P30EY006360 (NJ)

### Author Declarations

This cross-sectional study was determined exempt from formal review by the Emory University Institutional Review Board

