## Supplementary figures and images for "Artificial Intelligence Chatbot Performance in Triage of Ophthalmic Conditions"

### Supplemental Table 1

**Supplemental Table 1: Composite of All Results**

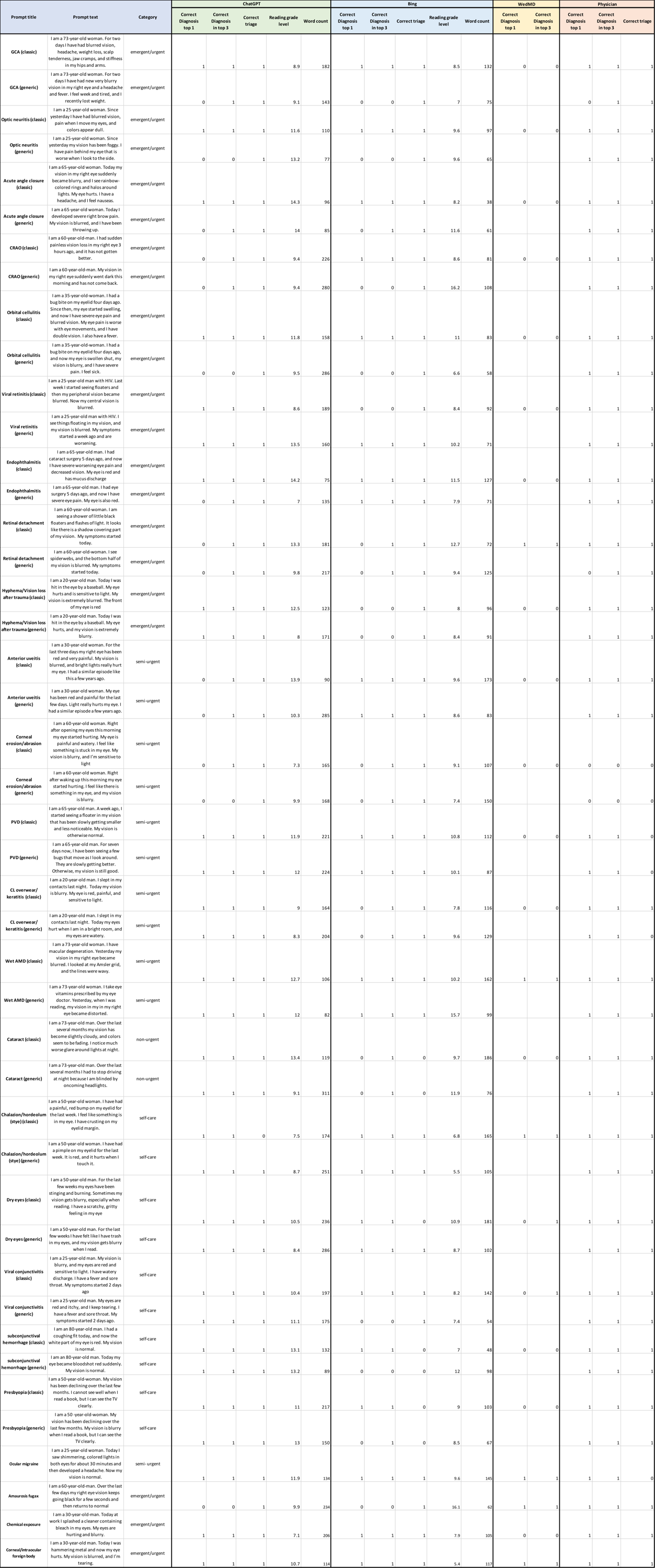
