## Supplemental Table 2 for "Artificial Intelligence Chatbot Performance in Triage of Ophthalmic Conditions"

S**upplemental Table 2**

|  | **Vignette Diagnosis** | **Prompt Language** | **Gross Inaccuracies** |
| --- | --- | --- | --- |
| **Bing** | Acute angle closure | Generic | Lists **macular degeneration** as a top diagnosis in the differential for severe right brow pain, blurred vision, vomiting |
|  | Corneal erosion | Generic | Lists **refractive errors** as top diagnosis in differential for painful eye and blurry vision |
|  | Wet AMD | Generic | Includes **juvenile macular dystrophy, keratoconus, astigmatism, conjunctivitis** for painless acute change in vision |
|  | Presbyopia | Generic | Lists **myopia** a top diagnosis for difficulty with reading, but intact distance vision |
|  | Cataract | Classic | Lists **"inflammation in your eye"** in differential and explains it as "white blood cells rush to contain the swelling, they can cause cloudy vision and other symptoms" |
|  | Viral conjunctivitis | Classic | When asked for references, provides organic remedies (**honey to eye**) that could be harmful |
| **WebMD** | Optic neuritis | n/a | Lists **cataract** in top 3 |
|  | Acute angle closure |  |  |
|  | CRAO | n/a | Lists **lazy eye** as a top diagnosis |
|  | Endophthalmitis | n/a | Lists **subconjunctival hemorrhage** as the top diagnosis |
|  | Hyphema/Vision loss after trauma |  |  |
|  | Anterior uveitis |  |  |
|  | Corneal erosion |  |  |
|  | Dry eyes |  |  |
|  | Viral conjunctivitis |  |  |
|  | Chemical exposure |  |  |
|  | CL overwear | n/a | Lists **subconjunctival hemorrhage** in top 3 |
|  | K foreign body |  |  |
| **Physician** | none | | |
| **ChatGPT** | none | | |

**Supplemental Table 2: Gross Inaccuracies:** Responses by Bing and WebMD included multiple gross inaccuracies, while responses by physicians and ChatGPT featured no gross inaccuracies.
